# ‘Isn’t social prescribing what social workers have been doing forever?’: UK social worker perspectives on social prescribing and professional boundaries

**DOI:** 10.64898/2026.04.24.26351583

**Authors:** Caroline White, Elizabeth Price, Liz Walker, Jo Bell, Lisa Revell

**Affiliations:** University of Hull

**Keywords:** Social Work, Social prescribing, Professional Boundaries, Interprofessional Working, health, social care policy

## Abstract

Social prescribing has assumed increasing dominance in policy and practice internationally, including in the UK, where it has an increasing role in addressing social needs such as isolation, and social determinants of ill-health. Although General Practitioners are perceived as key referral sources, social workers in one locality were found to play a significant role in referral. This suggests that the social work role in this context has been under-recognised and under-explored.

This study sought to explore social workers’ perceptions and experiences of social prescribing through an online survey conducted from January – June 2022. All UK social workers were eligible to participate, regardless of whether they had made referrals.

Responses (105) were collected from all UK nations. Data was analysed using inductive thematic analysis. Four key themes were generated: contended and contested boundaries; complementary spaces; delineated spaces of simplicity and complexity; social work under threat.

Participants recognised that social prescribing could provide valuable client support and could be a useful resource for social workers. However, they also expressed concerns about overlapping professional boundaries and the potential for social prescribing to encroach on social work, perceiving it as most appropriate for the delivery of support to those with ‘low-level’ needs.

## Introduction and Background

Although there is international interest in social prescribing (Morse et al. 2022; Scarpetti et al., 2024) there has been particularly significant and rapid growth in the UK. Recognising variation in definitions (Morse et al. 2022), Muhl et al. (2023, 9) have proposed an internationally relevant and acceptable definition of social prescribing as:

> *A means for trusted individuals in clinical and community settings to identify that a person has non-medical, health-related social needs and to subsequently connect them to non-clinical supports and services within the community by co-producing a social prescription—a non-medical prescription, to improve health and well-being and to strengthen community connections*.

In the UK, there has been a strong emphasis on social prescribing delivery supported by Link Workers (LWs) who are charged with enabling people to focus on the things that matter to them, and connecting them with community groups and agencies for practical and emotional support (NHSE/NHSI, 2020). However, it has been emphasised that social prescribing should be understood as *a pathway with many interacting elements* (Husk et al. 2020, 319), involving multiple individuals and relationships; this process includes inward referral/signposting; work undertaken by LWs and individuals to identify goals, needs and potential approaches to meeting these; onward direction to, and support from, the Voluntary and Community Sector (VCS) (Carnes et al. 2017; Husk et al. 2020; White et al. 2022). There is considerable heterogeneity within UK social prescribing. Some services are generic and open to a wide sector of the community, others have a narrower focus, in which eligibility is determined by age or specific health conditions (for example, Wildman et al, 2019) or outcomes sought, such as a reduction in loneliness and isolation (for example, Foster et al. 2021; Holding et al. 2022), with further variation in the duration of LW support and whether it is located in community or clinical settings (Keily et al. 2022). Further, some projects focus on specific places and the potential value of immersion and activity in green, blue and cultural spaces (Alejandre et al. 2023; Fixsen and Barrett 2022; Gorenburg et al. 2023). Across all UK devolved jurisdictions social prescribing has received policy attention (Department of Health, 2021, 2024; Fixsen and Barrett, 2022; Scottish Government 2012; Welsh Government, 2024). In England it has assumed increasing dominance within NHS policy, where it has been heralded as an approach to address the challenges and demands within primary care, loneliness and the growing pressures in mental health services (Department for Digital, Culture, Media and Sport (2018); NHS England 2016, 2019). Social prescribing has been a focus of the NHS Long Term Plan and primary care policy, and substantial investment has been made, with policy directives that all Primary Care Networks (PCNs) must provide access to social prescribing (NHS England 2016, 2019). This may be delivered by Link Workers employed by the PCN or commissioned from other agencies, often within the voluntary and community sector (Westlake et al. 2022). Thus, social prescribing spans the statutory and voluntary sectors, medical and social spheres, represents diverse ways of working, and aims to meet both policy goals (reducing pressures on general practice and diverting people with non-medical needs), and personal goals in respect of individual health and wellbeing. The use of social prescribing to address myriad goals has been termed by Pot (2025) as a *hyper-solution*, highlighting these diffuse and varied aims.

### The evidence base for social prescribing

Despite the strong policy focus on social prescribing, the evidence for its effectiveness is unclear, and methodological weaknesses in studies have been highlighted (Bickerdike et al. 2017; Pescheny et al. 2019) A systematic review published in 2017 concluded there was ‘little convincing evidence for either effectiveness or value for money’, yet ‘most evaluations have presented positive conclusions, generating a momentum for social prescribing that does not appear warranted’ (Bickerdike et al. 2017, 15). A more recent review (Kiely et al. 2022) also concluded there remains a lack of robust evidence for the effectiveness of social prescribing. Qualitative studies have provided evidence of benefits and change at an individual level, including improvements in well-being, management of health conditions, increased social connections and reductions in isolation, although benefits do not appear to be sustained over time for all (Carnes et al. 2017; Foster et al. 2021; Pescheny et al. 2019;). Benefits may also be unevenly distributed, as evidence suggests that areas of greatest deprivation have lower levels of LW employment, and with barriers to accessing community services reported for those least able to afford costs of travel and attendance, challenging the suggestion that social prescribing may help reduce inequalities (Hazeldine et al. 2021; Husk et al. 2020; Wilding et al. 2024; Wildman et al. 2019).

### Social prescribing and social work

Social prescribing policy and research have placed a strong emphasis on ‘the social’, and the solutions that social prescribing may offer in respect of complex *social* problems, forging *social* connections, addressing *social* needs, *social* isolation, *social* capital and *social* determinants of health. However, despite their focus on social aspects of wellbeing, social workers’ perspectives remain largely absent from research (see, however, White et al. 2022) and there has been little attention to the potential links between social work and social prescribing, although in some countries social workers appear to undertake the LW role (Morse et al. 2022; Scarpetti et al. 2024). Instead, the policy gaze is firmly fixed on social prescribing in the context of primary care, although The Local Government Association (2016) highlighted the role of Local Authorities in respect of social prescribing, which includes delivering services to which LWs may refer people and commissioning/delivering social prescribing. Social workers within one geographic area were found to be key sources of referral to a VCS-based social prescribing service, substantially outnumbering referrals from primary care, and reported valuing LWs’ roles in information provision, supporting clients on initial visits to agencies and groups, and providing support when the social work role ended (White et al. 2022), suggesting that despite a lack of policy directives, social workers may be establishing links with social prescribing services.

### Study aims

This study aimed to explore UK social workers’ understanding and experiences of social prescribing. This included considering whether, and how, they access social prescribing support for their clients; their reasons for doing so; their perceptions of social prescribing and its value (positive or negative) both for social workers and those they support.

## Method

### Sample and procedures

Social workers throughout the UK were eligible to participate, whether or not they had access to and/or experience of social prescribing. This approach was intended to capture both informed experiential accounts and the perspectives of practitioners with limited awareness of social prescribing, reflecting variation in knowledge and access across practice settings.

Online survey methodology was adopted as an approach which supports the exploration of under-researched areas, and which is untethered from temporal and spatial demands, enabling participants to respond at their convenience (Braun et al. 2020), helpful in the context of time demands on practitioners, and allowing wide geographic coverage. While surveys are typically perceived as a means of collecting quantitative data, they can also provide rich qualitative data (Braun et al. 2020). The survey captured demographic data about participants’ roles and geographic location and included quantitative and qualitative questions to explore whether participants have a social prescribing service in their local area; whether they have referred/signposted clients, and for what reasons; reasons for not referring; whether and how they perceive that social prescribing helps clients and social workers. This paper explores participants’ qualitative responses.

The research was publicised via social media, primarily through a project Twitter site (X - @SWSPstudy). Regular posts were shared with links to the survey; information was also shared with social work colleagues (in practice and education), social work students and teaching partnerships known to the research team. The survey was conducted from January – June 2022. All members of the research team were based in a university social work department, with professional backgrounds in social work and/or research in social work/psychology.

### Data analysis

Thematic analysis as described by Braun and Clarke (2006) was employed, as an established and well-defined approach to identifying and analysing patterns within qualitative data. An inductive approach to the analysis was undertaken with themes developed through close engagement with the data. The completed surveys were initially read by members of the research team, ensuring familiarisation with the data. This close reading supported the development of codes which were used to generate an initial thematic framework which was refined through successive reflection and discussion of the data in an iterative process. These themes were named to capture key meanings, with quotes employed to further illustrate thematic meanings.

### Ethical considerations

Approval for the study was provided by the University of Hull Faculty of Health Sciences Research Ethics Committee. The survey was completed anonymously, and participants were asked to give no identifying information. Given the anonymity of responses, completion of the survey was taken as consent.

## Results

105 completed surveys were returned.

### Participant characteristics

Participants were geographically diverse, representing all devolved nations of the UK, and all English regions. Yorkshire and Humberside had the highest representation (35% of respondents).

78% of participants were registered social workers. Participant job titles included social worker; Principal Social Worker; Approved Mental Health Practitioner; team manager/head of service; student social workers and apprentices also participated. The majority (74.5%) were employed by a Local Authority or Council; the remaining 25.5% worked within settings which included the VCS, the NHS, regulatory bodies, and charities. Participants worked across a wide range of social work services and with client groups from all stages of the life course.

Varying levels of knowledge and experience of social prescribing were reported by participants. A substantial proportion (35.2%) did not know whether a service existed in their locality. This limited both their engagement and their ability to evaluate its potential worth; some shared negative or positive perceptions, which are included in the findings below, while others stated they could not judge its value, reflecting their limited knowledge. Some indicated they would direct clients to social prescribing if they could. Of the 60 participants who were aware there was a social prescribing service in their locality, 28 (46.7%) said they had made referrals/signposted, while 32 (53.3%) had not.

Reasons for non-referral included fundamental criticisms of social prescribing, and having limited knowledge or opportunities to refer, although others perceived value in social prescribing; among those who had not referred there were mixed views about whether they thought they would direct clients to social prescribing in the future.

Through the thematic analysis four themes were generated, reflecting different understandings of professional roles, boundaries, and responsibility for addressing social need:

- Contended and contested professional boundaries: *‘it’s what we do’*
- Social work and social prescribing as complementary spaces: *‘this can take the pressure off us’*.
- Delineated spaces of simplicity and complexity: *‘complex issues that can’t be fixed with a social prescription’*
- Social work under threat: *‘why are we not trying to reclaim our social work roots’?*

### Contended and contested boundaries: ‘it’s what we do’

Connecting clients to community groups and resources (a key social prescribing role) was, for some participants, a core aspectLof the social work role, identity and training, and an established and regular element of social work practice:

> *As a social worker this is what we do all the time, we are considering the health and wellbeing of the person and looking to see what is out there to support their individual needs (SW82)*.
>
> *Isn’t social prescribing what social workers have been doing forever? (SW12)*.
>
> For others, social prescribing appeared akin to a bygone era of social work, which had been eroded:
>
> *Social prescribing is social work like it used to be, when you had time to work with people rather than just signpost or commission a package of care (SW08)*.

There was a perceived overlap between social work, social prescribing and other sources of support such as local area coordination and participation teams. This suggests that the ‘social space’ may be both fragmented and crowded, with potential for confusion among practitioners and the public alike. Further, understanding of social work among LWs was also criticised and reportedly led to ‘inappropriate referrals’, and disjointed referral pathways for the public:

> *The social prescribing service refers into adult social care a lot, most often inappropriately. We then refer to our local area coordinators to support the individual to access the universal services they’re after, which the social prescriber could have done (SW14)*.

Some participants contrasted the support provided by social workers with that provided by LWs. These accounts framed social work as providing superior support - *a more nuanced response (SW29)* - in contrast to social prescribing which appeared to be perceived as more limited in scope, and removed from the professional expertise and infrastructure of social work:

> *As far as I can see, social prescribing takes one element of social work, removes it from the wider field of experience and expertise, the networks of professional peer support, continuing professional development, reflective supervision, the values and principles of social work, and uses it within a medical model (SW15)*.

The above respondent further articulated their perception of the differences between these two areas of practice in respect of insight, skills and expertise: *It’s like asking astronomers if they could benefit from astrology (SW15)*.

Participants were critical both of the concept of social prescribing, which was perceived as a medicalised approach, and the language of prescribing in the context of social support:

> *There is a fundamental problem in using the term social prescribing to describe what is essentially a community support. Prescribing is a medical term. Important not to medicalise relationship based personal, family and community support (SW13)*.

However, others considered some clients to be more comfortable approaching practitioners within health settings and was more acceptable than social work support for some, and perceived that the language and authority of a prescription might lend weight to recommendations, encourage participation,

> *It is often a good way to engage hard to reach customers, as a lot of the time they don’t want to speak with social workers, but will speak to Social Prescribers (SW56)*.

The perception that social prescribing functions belonged firmly within social work were often aligned with very negative, sometimes hostile, attitudes to social prescribing. However, some appeared to have taken a pragmatic approach in which, whilst critical of social prescribing or recognising that, historically, social workers may have had a stronger role in engaging with community agencies, they had themselves made referrals to social prescribing services.

### Social work and social prescribing as complementary spaces: ‘this can take the pressure off us’

While some participants perceived social prescribing as a ‘poor relation’ or usurper which both duplicated and simplified the social work role, others perceived greater complementarity between them. These participants perceived that social prescribing could offer useful support when individuals do not meet eligibility criteria for social work support, or when social work involvement ends:

> *I have referred three separate people who have had moderate health and social issues. Social prescribing for two was a useful step-down option to ensure they had access to ongoing support which would be tailored to their needs. With another family it was because the parent was isolated and had volunteered before, expressing an interest in doing so again (SW47)*.

Social prescribing was perceived as having a preventative role, helping prevent crises, and diverting people from social work through a reduced *intervention from LA, and focus[ing] less on formal support to reduce dependency on services* (SW91).

Some perceived a role for joint working and narrowing the boundary between social work and social prescribing in which each practitioners’ role, expertise and time could be harnessed to support individuals:

> *Where organisational constraints limit the time I as a social worker spend with a person, the social prescribing service can spend that time looking at areas of needs myself and the service user have identified and look to address those needs. I then maximise my time supporting the individual with their mental health* (SW07).

Social workers also appeared to recognise the knowledge that social prescribers may hold of locally available services which are a resource both for social workers and for clients:

> *In my role I have one opportunity to refer the person and having this service means help could be better pinpointed, not just from my immediate knowledge (SW58)*.

Participants were asked whether they knew what support those they had directed to social prescribing services had received. Frequently they did not appear to know whether people attended and what (if any) support or benefits were received, although a small number reported receiving positive feedback. This often reflected their own relatively short-term, time-boundaried roles, in which they did not usually receive feedback from clients or LWs. Such information, if available, may inform social worker perceptions of social prescribing, and their understanding of its value and impact (positive or negative) and potential limitations.

The first two themes contrast, as they present critical (theme one) and more favourable (theme two) perceptions of social prescribing. However, some participants expressed opinions which suggested mixed views, perceptions and experiences. For example, in the quotes below, social workers who had referred to social prescribing noted variation in the quality of social prescribing services and support, and uneven benefits, in which the main advantages may be felt by social workers:

> *Some areas are run much better than others and some workers are more knowledgeable than others. I wouldn’t refer someone with ‘appointment fatigue’ unless I was absolutely sure that the service was good and that they would be matched with a good worker (SW97)*.
>
> *It helps me, however I’m not sure it helps the service user. It’s yet another service/organisation that’s getting involved (SW98)*.

### Delineated spaces of simplicity and complexity: ‘complex issues that can’t be fixed with a social prescription’

Social workers perceived a very clear delineation between the roles and activities appropriate to social workers and LWs, based on the complexity and depth of support individuals required. Social prescribing was described as offering ‘low-level’ support when individuals had ‘low-level’ needs, although these terms were not explicitly defined. Examples of ‘low-level’ support included activities such as ‘hand holding’, signposting to community resources, help completing forms and accompanying to appointments:

> *I think those with needs that could be addressed more informally would benefit. Those with higher care and support needs who require formal support would not (SW86)*.

Social prescribing was also perceived to have a role in helping address loneliness and isolation, financial problems and debt, emotions such as ‘low mood’ and anxiety, and providing interim support while awaiting the involvement of statutory services:

> *We work with a lot of parents who are socially isolated with low level MH issues who are unable to access MH services or have a long wait (SW41)*.

In contrast, social workers identified their own roles as supporting clients with ‘complex’ issues. For some, social prescribing appeared insufficient to address such issues, and this was given by some as a reason for not referring:

> *Because the people who use our services have complex situations that require a fuller understanding and are rarely resolved by a one-dimensional signposting response (SW29)*.

Others, however appeared to perceive potential value in working with LWs, with roles boundaried by narratives of simplicity and complexity, in which it was considered that the LW could support with issues which might not meet the thresholds for social work support:

> *I anticipate it may be a quicker way to access low level support and would then free up time for registered social workers to respond to safeguarding concerns etc in a more timely manner (SW38)*.

This delineation of roles sometimes appeared to be a further source of criticism of social prescribing (suggesting this is an unskilled area of work) or could reflect a sense of complementarity in which both social workers and LWs could contribute to individuals’ support, and with social prescribing appearing a valuable resource to undertake tasks outside the scope of social work, and extending the options available both to social workers and clients:

> *In an age where services are retreating its useful to have somewhere else they can go*
> (SW47).

### Social work under threat: ‘why are we not trying to retain/reclaim our social work roots’?

Some participants articulated a clear sense of social work as under threat from a political and commissioning system which did not value social work and social work skills, and in which health services and medicalised approaches were perceived as more aligned with public and political support. Social work was described as underfunded, with the social work role eroded and often reduced to core functions and with social workers experiencing substantial time pressures:

> *It’s an ongoing struggle within social work that funding has been further and further stripped back over the years, reducing social work just to organising packages of care, rather than holistically enabling people to access their community and living fulfilled lives (SW15)*.

In contrast to the view that social prescribing could complement social work, some perceived it was undermining social work skills and values, necessitating a need to resist the erosion of the social work profession and to reclaim their social work roots:

> *If you are a social worker you should not endorse prescribing or signposting. Social prescribing is not contributing to social work it is leading to the loss of our core skills and values. Social work needs to re engage with its radical roots and ditch the flawed agenda of social prescribing (SW11)*.

However, this participant cautioned that pressures of work could mean that social workers are too over-burdened to recognise the threats to their roles, believing that many experience an unquestioning and welcome relief at the availability of a form of support which they are not themselves in a position to provide:

> *More than ever, the world is now viewed through a medical lens and social workers are likely to be too busy to recognise this. I fear your survey will be skewed by responses from SWs who haven’t had time and space to think through the implications of either the medicalisation or simplification presented by SP and who will be relieved by the offer of resource and cash available to respond to need without critical thought on how it’s being offered (SW29)*.

## Discussion

This research sought to capture social workers’ views about social prescribing which, although a key focus of recent health policy and research, has not, to date, incorporated a social work perspective. Through engaging with social workers, significant questions were raised about the nature and scope of support for those with ‘social needs’, the practitioner roles best suited to meeting these, and the training and resources required for those delivering a new ‘brand’ of support within an ever-pressurised social space. Further, in exploring this new provision, the research identifies a sense of social work as a profession under threat.

The data captures a range of experience, knowledge and perceptions, with participating social workers sharing a wide spectrum of viewpoints. At one end of this spectrum were those who could be considered *idealists and/or critics*. These participants expressed very negative and critical views of social prescribing which they appeared to perceive as providing a diluted and diminished form of social work, viewing it as a threat to social work roles and values. At the other end of the spectrum were those who could be characterised as *broadly supportive* of social prescribing, these appeared to take a more positive view, appearing to recognise some complementarity between the two roles. In this context, there was a willingness to refer clients onwards when their needs could not otherwise be met, or to engage in joint working with social prescribers, harnessing their respective skills, with social workers undertaking the more complex tasks for which they felt they had been educated and trained. There were also participants whose views were at the less extreme ends of this spectrum, including those with mixed views. These included social workers who were critical, yet had referred people to LWs, appearing to adopt a pragmatic perspective, and others who were uncertain about the value of social prescribing. A high proportion of participants had limited knowledge, awareness and experiences of social prescribing. This is both a significant finding, as well as a study limitation.

Participants who were critical of social prescribing often expressed the most trenchant and clearly articulated perspectives, identifying it as an inappropriate response to individuals’ needs, which encroached upon social workers’ roles and expertise, in an over-medicalised context. Respondents who had directed clients to social prescribing often gave less strident responses, which lacked the detail or obvious enthusiasm which might have matched the views of their more critical counterparts. These less forcefully articulated views may reflect the observation that, as cases closed, they received little feedback about whether the person accessed, or benefitted from, support, as well as their own relatively short-term involvement and the relative newness of social prescribing. As experience and knowledge of social prescribing grows among social workers, they may develop stronger (positive or negative) opinions.

The social worker narrative drew a clear delineation between their own work, which was articulated as complex, in-depth and skilled, and the less complex, ‘low-level’ support which was perceived as appropriate to the LW role. This perspective appears to align with the policy rhetoric, which presents a contradictory and ambiguous narrative of both the simplicity and complexity within the LW role, while situating this as largely straightforward and requiring minimal professional skills. However, over successive policy iterations, there has been a creeping acknowledgement of the complex realities of social prescribing, although this is not fully acknowledged or addressed. Recent policy directives have pronounced that social prescribing is suitable for those with *low leve*l mental health needs, and people who are lonely, have long-term conditions, and for those who have *complex* social needs, which can be addressed through a *simple* care and support plan (NHS England 2023; NHS England/NHS Improvement 2020; our emphasis added). ‘Complex needs’ are not defined, but there is a glancing acknowledgement, that that those who come into the orbit of social prescribing may be in crisis, and that their experiences may include domestic violence, sexual abuse, self-harm and suicidal thoughts, all experiences which are far outside the sphere of the stated ‘low level’ needs. Further, these policies stipulate that the LW role is suitable for those whose lived or voluntary experience enables them to draw on personal experience and qualities, but with little evidence of in-depth training to complement these personal attributes and insights (NHS England, 2023), suggesting that empathy and awareness are perceived as sufficient to ensure appropriate support. This, arguably, reflects a process of de-professionalisation in which the social work role has both been absent from this policy development, and has increasingly been undertaken by other professions, most tellingly, in the context of this research, unqualified workers, consequently undermining social work as a regulated profession (Spolander et al. 2014). LWs themselves have reported that they feel neither prepared nor adequately trained to support people with the in-depth support needs they encounter within their roles (Hazeldine et al. 2021; Rhodes and Bell, 2021;Wildman et al. 2019;), and some appeared to feel they were expected to address a gap which should have been provided by statutory services and were *acting as unqualified social or advocacy workers for service-users* (Holding et al. 2020, 1539), further indicative of a process of de-professionalisation.

These competing narratives of the simplicity/complexities of social work and social prescribing, and the shifting roles within both, raise important questions about how those who require assistance with the social elements of human experience and wellbeing are supported, and by whom. The statutory social work role has, over time, become increasingly focussed at either end of the safeguarding spectrum with support provided only to those with the greatest needs and those assuming the highest degree of ‘risk’ (evidenced by increasingly restrictive eligibility criteria), with the social work role focussed on assessment, rationing, and gatekeeping; this, inevitably, limits the nature of the support social workers provide and the populations they can work with (Jones, 2014; Lloyd et al., 2014; Pulman and Fenge, 2024). Within this landscape the community development role within social work has been largely overshadowed, with relationship-based practice being discouraged, although there is also evidence of social worker discretion and of going ‘underground’ to extend their roles when clients do not meet eligibility criteria or when formal involvement ‘should have’ ended (Jones 2014; Moth et al. 2024). In the context of the contracting statutory social work role, there is an apparent support gap which, it seems, social prescribing is, as identified above, in some instances, being used to address.

Recent ethnographic research, however, indicates that, as the population accessing social prescribing appears to be widening, there may also be pressures to change the approaches taken, which may, ironically, mirror those which have beset social work. These studies shine a harsh light on social prescribing practice in some areas, which appears to have become more target-driven and procedural, less flexible and responsive to individual need, with an increased requirement for individuals to assume greater responsibility for their own care, and with clients increasingly signposted onwards rather than receiving more active forms of support (Gibson et al. 2022; Griffiths et al. 2022; Westlake et al. 2024). This suggests that, just as the social work space has contracted to focus on those assessed as in greatest need, and on gatekeeping to manage demand, so too might social prescribing be increasingly subject to similar constraints. This raises questions about how people who have significant needs, but who do not meet the criteria for social work involvement, are supported (and the quality of that support), and whether those with less intense support needs, for whom social prescribing was previously envisaged as an effective approach, might be squeezed out of the social support space. How those who are ineligible for, or are moved onwards from statutory support, and who feel unable to manage and direct their care, fare in these increasingly time limited contexts, requires further exploration.

Our findings are mirrored in those of Bradley et al. (2025) who explored Occupational Therapists’ (OT) views and experiences of social prescribing. Participants perceived an overlap between the OT and LW roles, challenges to their professional identities, and believed they should remain responsible for more complex work. Others, like their social work counterparts, worked alongside LWs, and engaged with social prescribing for step-down support. Together, these findings suggest that the boundaries between social prescribing and professions such as social work and occupational therapy are porous, with overlap between professionals whose roles include support in respect of social and emotional needs and wellbeing, and those of LWs. These findings collectively signal the need for a conversation in which there is careful consideration of professional boundaries, the role it is reasonable to expect of social prescribing and the limits to these, while ensuring that social prescribing is not used as a substitute for the practice wisdom within established professions which have well-developed evidence bases, professional regulation, and long-standing professional training and development programmes. The current ambiguous policy landscape does little to inform this, through uncritically leaning on social prescribing as a solution to multiple individual, societal and practice issues (chiefly within primary care). This exemplifies the hypersolutionism described by Pot (2025) in which social prescribing is expected to respond to unrealistically diverse agendas, and ‘silver bullet thinking’ (Eccles, 2021), in which there is over-reliance on a single and simple policy solution to complex problems.

### Strengths and limitations

The online survey methodology enabled participation of geographically dispersed social workers, with a wide range of roles and experiences represented, within a neglected area of research. There was variation in the level of detail within participant responses; some provided in-depth answers, while others gave relatively little detail, and the survey methodology did not enable probing for additional information or when clarification was needed. As social prescribing becomes further established, and pressures on social work appear ongoing, it will be important to continue to explore the relationship between social work and social prescribing, supported by additional methodologies, such as focus groups and interviews to complement survey findings.

## Conclusions

Findings from the study highlight the complex, and often conflicting, perceptions of social workers in the context of social prescribing. While contemporary policy promotes social prescribing as a solution for addressing social and health needs, some social workers view it as a diluted version of their role which threatens their professional identity and is divorced from social work values, although others perceived greater complementarity between their roles or engaged with social prescribing through pragmatic recognition of its value in the context of their own time constrained roles and restrictive eligibility criteria.

The evolution of social prescribing, however, appears to mirror the challenges faced by social work itself, particularly in the context of austerity-driven policies. Just as social work has become more target-driven, focused on managing (high) risk and rationing services, there are signs that social prescribing may follow a similar path, potentially excluding some it was initially intended to help, with an apparently increasing focus on those requiring in-depth support. This raises concerns about the boundaries between the two roles, and whether social prescribing is being used to bridge the gaps left by increasingly constrained social work services. While social prescribing holds promise as a complementary support system, its implementation must be re-examined to ensure it does not compromise the integrity of social work (and other cognate professions) or contribute, ironically, to the further marginalisation of vulnerable populations. More in-depth training, clearer role definitions, and stronger feedback loops are essential to ensuring that social prescribing and social work can productively coexist and effectively address the increasingly wide spectrum of health-related social needs. We would argue that it is the wider social contexts within which social workers *and* social prescribing services operate that it is most important to recognise and respond to. Thus, we suggest that an increasing focus on the need for shifts in professional training and on the recognition and operationalisation of professional boundaries, whilst no doubt important might, paradoxically, only serve to limit social work’s sustained focus on resource-constrained service provision. In short, there should be a wider reflection on what constitutes an appropriate professional response to the marginalising cultures and practices which are an increasing feature of contemporary social life.

## Data Availability

Data produced in the present study is not available to other researchers.

## Acknowledgements

We would like to thank all social workers who shared their views and experiences, and the agencies and individuals who circulated information about the study to potential participants.

